# The Effect of Weather Pattern on the Second Wave of Coronavirus: A cross study between cold and tropical climates of France, Italy, Colombia, and Brazil

**DOI:** 10.1101/2021.12.28.21268496

**Authors:** Ahmed Islam

## Abstract

This study aims to explore and understand the common belief that COVID infection rate is highly dependent on either the outside temperature and/or the humidity. Thirty-six regions/states from two humid-tropical countries, namely Brazil and Colombia and two countries with temperate climate, France and Italy, are studied over the period of October to December. Daily outside temperature, relative humidity and hospitalization/cases are analyzed using Spearman’s correlation. The eighteen cold regions of France and Italy has seen an average drop in temperature from 10°C to 6°C and 17°C to 7°C, respectively, and France recorded an addition of 2.3 million cases, while Italy recorded an addition of 1.8 million cases. Outside temperature did not fluctuate much in tropical countries, but Brazil and Colombia added 4.17 million and 1.1 million cases, respectively. Köppen–Geiger classification showed the differences in weather pattern between the four countries, and the analysis showed that there is very weak correlation between either outside weather and/or relative humidity alone to the COVID-19 pandemic.

## 1. Introduction

Recent studies by different researchers show that weather temperature, humidity and precipitation may have largely contributed to the spread of influenzas and airborne viruses that are mediated through the means of aerosol droplets of different sizes. Human to human transmission of acute respiratory viruses, such like SARS-CoV-2, has turned into a widespread pandemic, with large fractions of infected patients suffering from acute respiratory distress syndrome (ARDS) and needing non-invasive and invasive mechanically ventilated interventions[1]. While the virus persisted throughout the year of 2020, hospitalizing thousands of patients all across the United States, many researchers claimed that the pattern of rise-fall-rise (winter-summer-winter) of the rate of daily infections indicates that the respiratory virus has a strong correlation with the seasonality, particularly with the changes in temperature, relative humidity (RH), absolute humidity (AH) and host behavior[2]–[4].

Because of the potential resemblance of typing of SARS-CoV-2, researchers have studied surrogate models to find out the survivability under different environmental settings. Typical healthcare environments with varying relative humidity (RH) but an ambient temperature (AT) at around 20C showed that potential surrogate virus types like transmissible gastroenteritis virus (TGEV) and mouse hepatitis virus (MHV) loose very small infectivity within a period of two days. Studies also indicated that TGEV and human coronavirus 229E survivability at low temperature and medium and low RH is rather enhanced.

A recent COVID-19 study [5] on droplet dynamics showed that the spreading and concentration of contaminated droplets’ have strong and significant correlation to weather temperature and humidity. Their numerical simulations of droplet spreading through coughing and sneezing has shown that at low temperatures (0°C) the spread of contaminated respiratory droplets would be quite wider and larger in spatial sense, compared to the spread of droplets at 20C to 40C. Similarly, at high RH (50% ∼ 90%), the contaminated droplets would thin out less compared to the spread at low RH (10% ∼ 30%). Therefore, in terms of temperature and relative humidity there is a strong correlation between high relative humidity at low temperature and the increased spread of concentrated and contaminated virus borne droplets. Another study [6] on the evaporation modelling of coughing droplets in high humid areas, where it was found that dry conditions enhance droplet travelling more efficiently than in wet conditions. The evaporation model study arrived in another major conclusion that smaller droplets are not affected by higher relative humidity (60% to 90%) compared to bigger droplets. Their final impression is that even though the evaporation model shows significant increase in evaporation rate with bigger droplets, the scarcity of study on the dilution and inactivation of small droplets in low humidity condition makes it difficult to assess the certainty of spreading and suspension of virus borne coughs and sneezes in different regions of the world. Iqbal et al. [7] and Bukhari et al. [8] concluded that coronavirus spread was faster in colder regions compared to warmer region and that there is close relationship between daylight hours, average temperature and risk of COVID infection rate. In different parts of the world, researchers found that there indeed positive correlation between COVID infection rate and humid climate. For instance, Pani et al. [9] found that along with temperature and weaker correlation with relative humidity, dew point and water vapor has positive correlation with COVID-19 in Singapore, a predominantly “hot and humid climate with abundant rainfall”. On the other hand, Takagi et al.[10] found negative association of temperature, pressure and UV with COVID-19 prevalence in Japan and exclaimed that the finding of no association of Covid-19 with climatic conditions in China [11] can be possibly argued. Both research papers were published based on the studies done in early period of COVID pandemic in specific geo locations (Chinese cities: Yao et al. [11], published in April 2020 and Japanese cities: Takagi et al. [10] published in August 2020). Similarly, a supportive study results from Japan showed that the epidemic growth has strong correlation to increase in daily temperature[12]. A very recent study done by Zhu et al. [13] looked across various regions in South America but concluded that among other factors, absolute humidity was highly negatively correlated to the COVID-19 spread. Across the 122 cities in China, Xie et al. [14] found that at certain threshold temperature of 3C, the mean temperature has positive linear relationship with infection cases and in Iran, humid provinces has higher rate of increase in infection rate and extreme dry regions have proved a reverse relationship[15]. Both in Brazil and Indonesia, Auler et al. [16] and Tosepu et al. [17] found that higher mean temperature and humidity has positive correlation in infection spreading which is in contrast to many other studies done in colder European and US regions[18]. Auler et al. [16] also reported that among the five Brazilian cities, Sao Paulo was the city with highest confirmed cases but with the lowest mean temperature and highest relative humidity. But with further statistical analysis they arrived at the conclusion that the disease transmission rate was favored by high temperature and relatively high humidity. Therefore, it can be assumed from their study that there is no strong correlation but rather several anomalies within a given region, and therefore a sole factor cannot be singled out to have strong impact on the increasing infection rate. In Victoria, Mexico [19] temperature was found to be negatively correlated to the spread of the infection and their study spanned from March 2020 till June 2020 but consequently did not include the sharp rise in infection rate of the second wave in other Mexican cities. Another study on tempered climate stated that tropical climate slows spreading of COVID-19 local transmission, and also reported to have negative association between temperature and local positive cases[20]. A case study based on New Jersey by Doğan et al. [21] produced results indicating that humidity has positive relationship and temperature has negative relationship to COVID-19 based on data collected and analyzed from late February to late July of 2020. They also pointed out that their study outcome is in contradiction to the study by Ahmadi et al. [15] in Iran, which stated that there exist strong correlation between COVID infection and humidity, temperature and wind. An associative study has explored the pathway of COVID-19 spread in Oslo Norway a little differently, where Menebo et al. [22] implied that sunny weather makes people come out of home and rainy weather makes people stay indoors, and hence warm climate triggers an increase in infection and spreading events. Many studies found strong temperature association based on low COVID cases in different countries, as pointed out by [23] and there remains the question as to what happened afterwards with regards to exponential global growth in infection and death inherently affecting different individual regions. Bashir et al. [24] indicated that scientific evidence does not support that warm weather would bring down the epidemic spread contrary to popular misbelief pointed out by many researchers [25] when compared to different influenza and COVID variants [26], [27]. In Spain[28], Iran[29] and in 50 US cities[25], studies conducted between February and March showed that there exists no correlation between weather variables and COVID-19, which contradicts to the other studies that found some correlation as discussed before. Even recent observations by Pan et al. [30] implicated that meteorological factors, including temperature, did not exhibit significant association and would not help in reducing COVID-19 transmission. Several other studies that studied the mixed combination of different climatological factors have either found unconvincing or very weak correlation to COVID transmission [31]–[33].

It is also relevant to mention that several studies have [11], [34]–[38]. On the other hand, many studies have confuted weather factors that were deemed strongly correlated to the rate of spread of the infection and argued the weaknesses of different studies[39]. Certain researchers pointed out that the factors like population density, emergency care and medical treatment, socio-economic conditions of different locations could be coupled with climatic factors and thus disassociating or considering outside temperature or humidity to be a single controlling factor would give false perception, conception and pretense on how SARS-CoV-2 spreads[23], [40].

In this study, our approach to understand and elaborate the difference in correlation between climatic conditions and the coronavirus transmission is based on a total of four countries, two countries that have relatively dry colder climates and two that have tropical humid climates during the period of October to December of 2020. In the later part of the paper, we would demonstrate, as many other research studies already pointed out, that a ***single*** climatic factor is not solely responsible for the spread of the coronavirus infection among different types of climate regions.

Part of the problem with statistical correlation is always related to the degree of uncertainty and the risk of over-confidence in statistical representation of the results. While many of the statistical studies are done with relatively low spread of infection (compare to the spread and infection rate of COVID during the summer in US) researchers publishing data based on the late winter (February to April) and Summer is not totally indicative of the link between climate and COVID infection. This became more apparent in our study where we found that the weather model and the rise in infection in cold climatic regions (for instance in Italy, France) is totally opposite to tropical regions (like Brazil and Colombia) during the months of November and December. We acknowledge that climatic factors like outside temperature and humidity alone cannot predict viral transmissibility and the spread of the SARS-CoV-2 infection, rather physiological factors through means of aerosol and infected droplets causing membranous fusion and are found to be dependent on wet-bulb temperature which in turn is a function of indoor/outdoor room temperature, absolute and relative humidity, as investigated by JD Runkle et al. [41] and Dougherty [42], are the active route of transmission for the virus. What is more important to understand is, measure of social distance, mask mandates and part of governing policy regulation including lockdowns are key factors that dictate the rate of infections, not the weather as believed by many including policy makers.

## 2. Methods

### 2.1 Data Collection and Validation

For this study, weather data is collected from Integrated Surface Database (ISD) from NOAA’s National Climatic Data Center (NCDC) [43]. The ISD data from more than 20,000 stations worldwide and consists of different weather identifying subsets including, but not limited to, World Meteorological Organization(WMO), Weather Bureau Army Navy (WBAN), Climate Reference Network (CRN), Federal Aviation Administration (FAA), Automated Surface Observing System (ASOS), and Automated Weather Observing System (AWOS) [44]. With extensive hourly and daily data including air temperature, dew point temperature, maximum and minimum recorded temperatures for the day, and wind speed, this study used the ISD provided data for the entire year of 2020. Several sources are used to collect the daily infection data for each of the four countries: Brazil[45], Italy[46], France[47], and Colombia [48]. Datasets have been crosschecked and validated with John Hopkins Coronavirus Resource Center [49], The New York Times [50], Google [51] and Microsoft Bing[52].

### 2.2 Calculation based on Longitude, Latitude and of Relative Humidity (%RH)

An extensive algorithm has been developed in MATLAB to study the spreading of COVID infection in two tropical countries Brazil and Colombia, as well as two temperate climate countries, Italy, and France. Regions/cities with highest reported cases in each country were picked and close proximal stations were identified using the longitude and latitude data, while cross checked with the Hourly/Sub-Hourly Observational Data Map [53]. The location and daily COVID cases/hospitalization information were very critical, since some of the hourly data were not available for some of the stations and some of the COVID data had lapses (unreported, erroneous or skipped reporting). Therefore, careful consideration has been made to locate correct WMO/WBAN stations within the given latitude and longitude combinations for each of the 36 regions/states and the weather data were accurately collected and matched with the COVID datasets using the MATLAB algorithm. Using the outside temperature and dewpoint temperature, the Relative Humidity (%RH) was calculated using the following relationship:

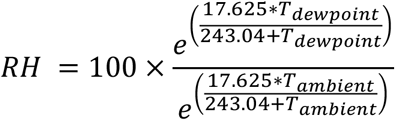

### 2.3 Analysis

Weather data and infection rate (in some countries reported as number of cases with Hospitalization) are analyzed from October 1^st^ to December 31^st^. Spearman correlation coefficients with bivariate, two-tailed analysis stating 95% confidence interval are also reported for each region where the infection and the weather patterns are plotted. (See **Supplemental Information** for Temperature and Relative Humidity data plotted against highest recorded infection/hospitalization cases for a total of thirty six regions of each of the four countries.)

## 3. Results and Discussion

Since October 1^st^, the outside air temperature started to fall in France and Italy, but a similar pattern was not observed in the two tropical countries considered, namely Brazil and Colombia. Because of the geolocation of Colombia, which is very close to the equator line, the seven-day averaged temperature did not deviate much. For instance, in between October to December, Bogota observed temperature change from 13°C to 11.5°C; Cartagena observed 28°C to 27°C. Except Tolima, all other regions reported very weak to almost no correlation coefficient (r_T_<0.30) in between air temperature and daily reported cases. Throughout Colombia, the weather classified by **Köppen–Geiger** moves from tropical savanna climate (Aw/As) to tropical monsoon (Am) to tropical rainforest climate (Af) the further the reference location moves from the equator line. While Bogotá and Antioquia weathers are classified as oceanic climate (Cfb) and warm tropical (Af) respectively, with outside temperature steadied at 13°C and 27°C and relative humidity ranging well within RH∼ 72% to 80%, the infection rate kept a steady record regardless of the outside air temperature and relative humidity. Considering only relative humidity (RH), for the highest recording nine departments of Colombia, shows no correlation (r_%RH_ <0.20), even though the relative humidity for Valle del Cauca, Norte de Santander, Huila, and Tolima were within the range of RH< 71% and Cartagena, Santander and Atlantico had steady record of RH > 80%. Thus, in both cases of air temperature and relative humidity, throughout Colombia there was very little correlation between weather and the spread of the second wave of COVID-19 infection through the months of October till December.

In Brazil, a widely varying climate is observed across all the regions, and in between October 1^st^ and December 31^st^, except for Santa Catarina and Rio Grande do Sul, the temperature varied in between 35°C to 25°C. Outside temperature for Santa Catarina and Rio Grande do Sul distributed between 25°C and 15°C, and the **Köppen–Geiger** classification for both states is considered as Aw (tropical savanna climate) and Cwa (dry-winter humid subtropical climate). In both states from mid-October to the end of December, the recorded infection/hospitalization rose from average of 2000 to 5000 and the Spearman correlation coefficient indicated a no correlation (r_T, Santa Catarina_ ∼ 0.18, *p-value*>0.05) to weak correlation (r_T, Rio Grande do Sul_ ∼ 0.41, *p-value*<0.05). Rio de Janeiro, Goias, and Ceara, all within the Aw (tropical savanna climate with dry-winter characteristics) has experienced an average of 3700 daily cases with little deviation from mean. With Ceará having hot-overall weather throughout region, the COVID infection kept spreading when the weather was within the overall dryer climate. On the other hand, in Goiás, the second wave was within the rainy season (October-April), but the infection rate soared throughout the time. Relative humidity for both Ceará and Goiás fluctuated between 40% to 60% while in Rio de Janeiro the average RH ∼ 80%, but calculated correlation coefficient were still very insignificant (r_%RH, Rio de Janeiro, Goiás, Ceará_ ∼ -0.07, -0.23, -0.03, *p-value*>0.05). In Rio de Janeiro, weather moved from spring to hot-humid summer from October to December, but infection record remained within 3700 cases every day. In no correlation (r_T, Rio de Janeiro, Goiás, Ceará_ ∼ -0.11, 0.21, -0.02, *p-value*>0.05) between the temperatures and the infection cases, thus the spread of COVID infection has very little correlation within this study period for Brazil.

**Figure 1:**
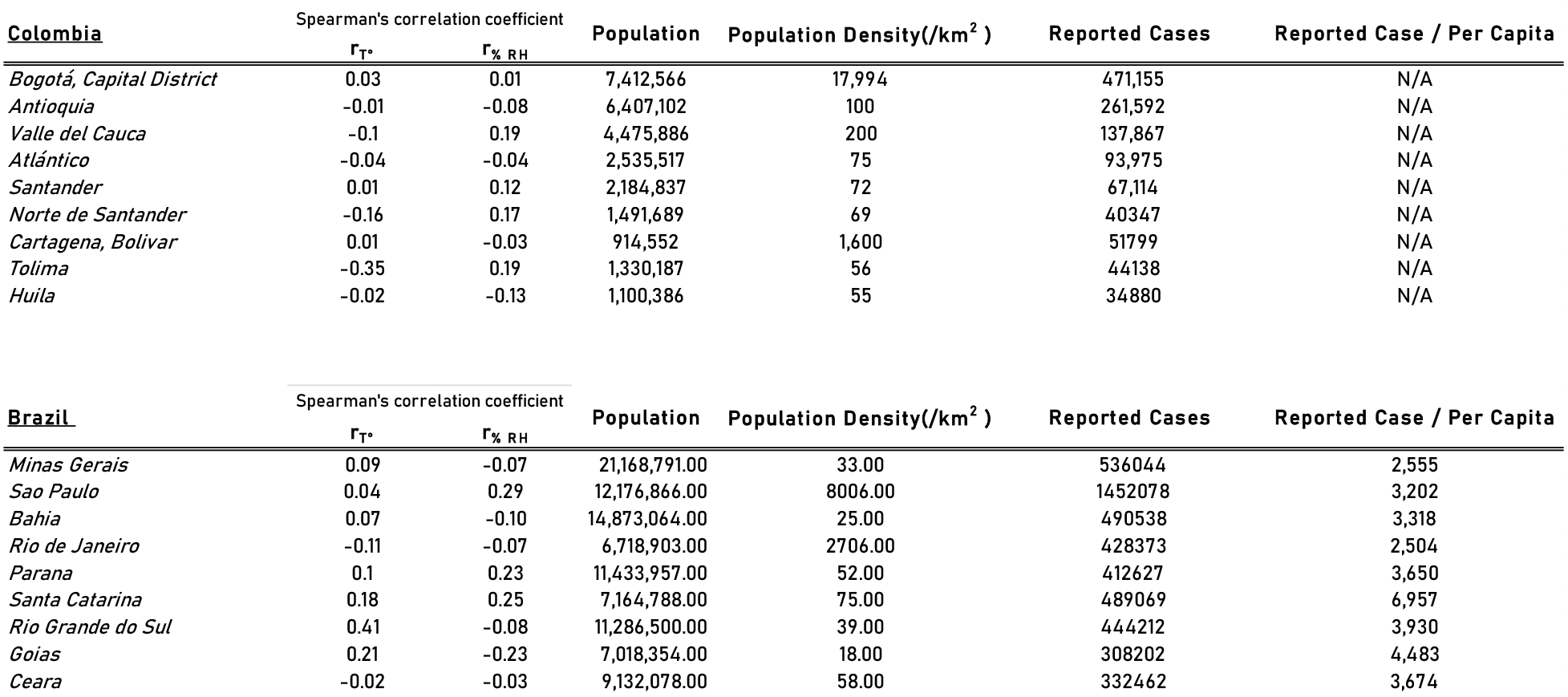
Spearman’s Correlation for Temperature and Relative Humidity vs Nine states/regions with highest COVID infection by the end of December 31^st^, 2020 of a) Top: Colombia and b) Bottom: Brazil

**Figure 2:**
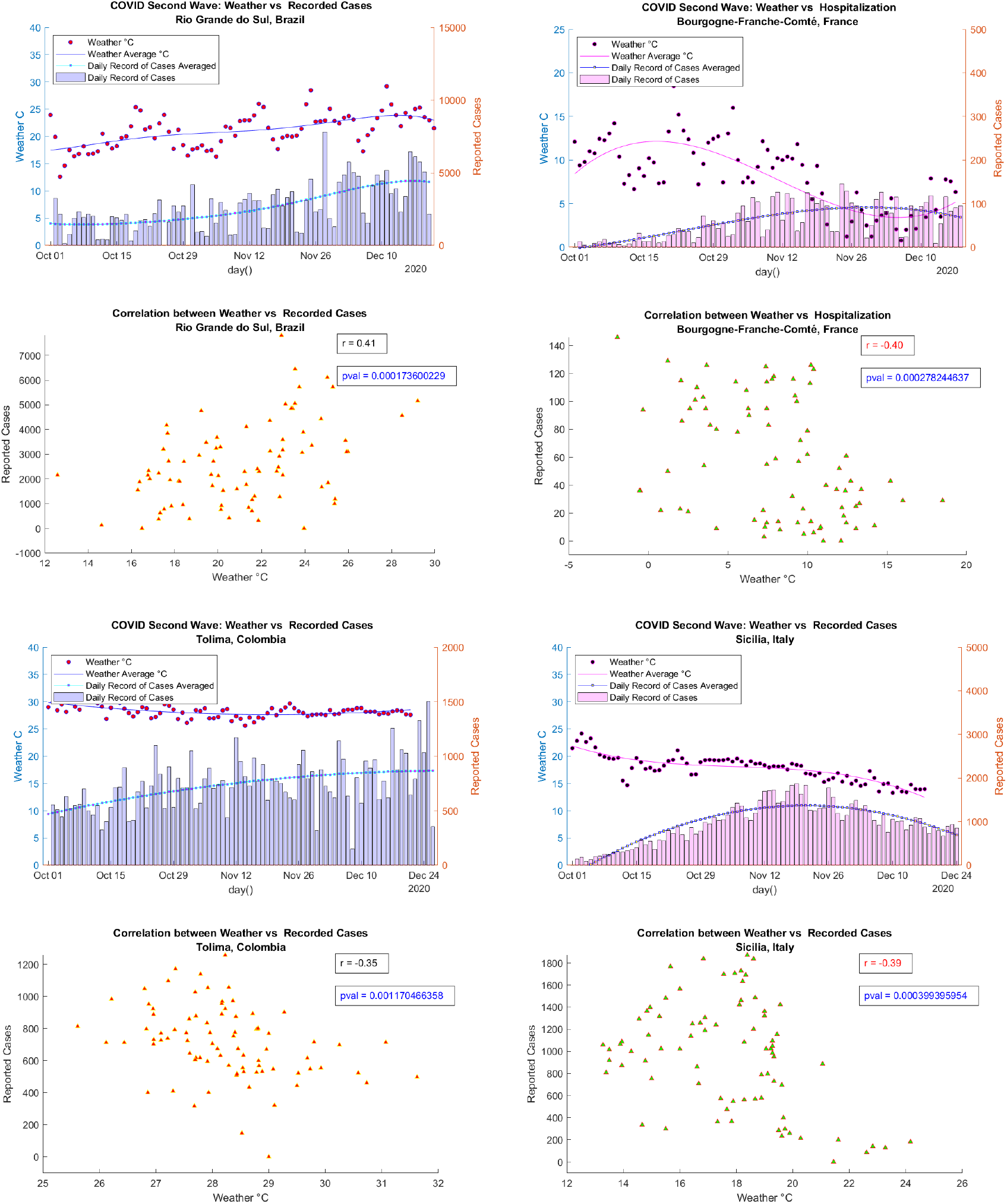
Comparison between recorded outside temperature patterns and the COVID infection of four different regions for two different climate types: *Rio Grande do Sul, Brazil* (r=0.41); *Bourgogne-Franche-Comte, France* (r=-0.40); *Tolima, Colombia* (r=-0.35); *Sicilia, Italy* (r=-0.39)

For colder climates, by the end of the year 2020, France and Italy recorded a total of 2.64 million (3.94% of the total population) and 2.14 million cases (3.55% of the total population), respectively. On the other hand, Colombia had an estimated of 1.67 million cases (3.28% of the total population) and Brazil had 7.72 million cases of infected people (3.62% of the total population). All four countries considered in this study have varying climate patterns, as shown in Figure 3, even though the proportions of people infected with the virus by the end of 2020 are approximately very similar. From the very beginning of October till mid-December, nine of the regions with the most recorded COVID cases in France, observed a constant fall in temperature, with an average shift of mean temperature from 13°C to 7°C. This observation is reflected in the correlation coefficient, especially in Auvergne-Rhone-Alpes (ARA), Grand Est (GE), and Bourgogne-Franche-Comte (BFC) regions, the Spearman’s correlation reported within a range of (r_T,ARA,GE,BFC_ ∼ -0.32, -0.56, -0.40, *p-value<*0.001). COVID cases, except in the case of Grand Est, is not significantly correlated to the reported related humidity.

**Figure 3:**
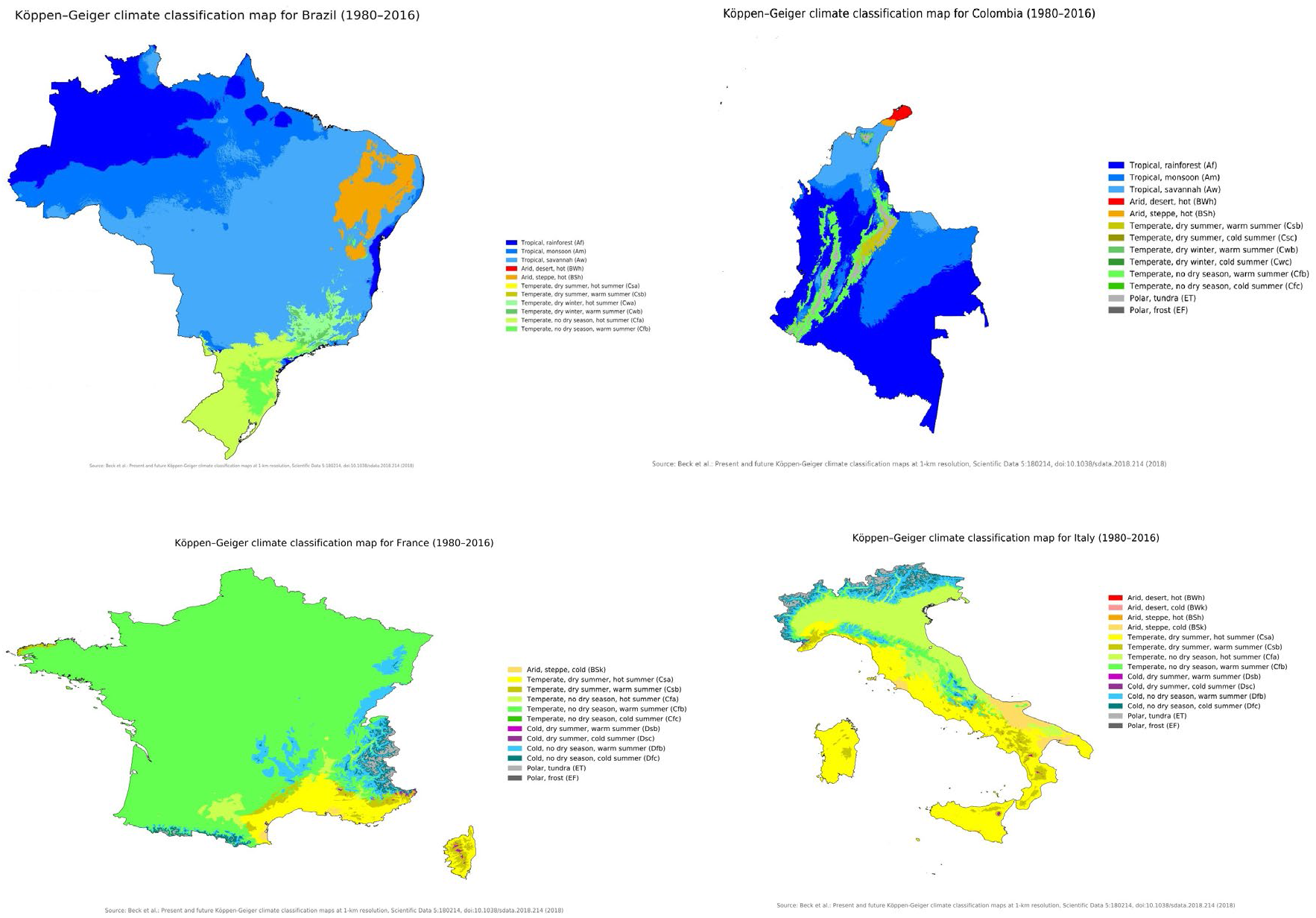
**Köppen–Geiger** classification of four countries: Top: Brazil (left), Colombia (right) Bottom: France (left), Italy (right) [54]–[58]

**Figure 4:**
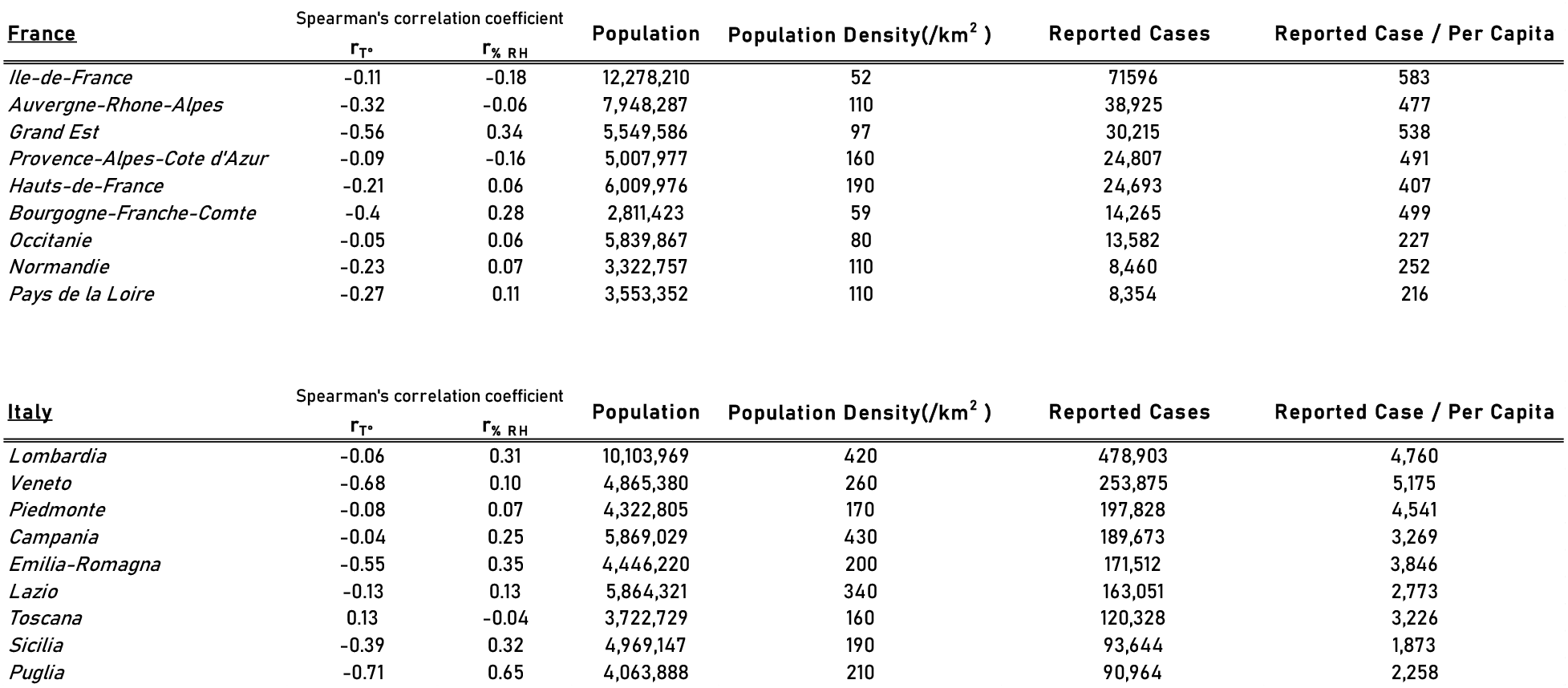
Spearman’s Correlation for Temperature and Relative Humidity vs Nine states/regions with highest COVID infection by the end of December 31^st^, 2020 of a) Top: France and b) Bottom: Italy

The COVID second wave in Italy has indicated an overall strong to moderate correlation in regions like Veneto (r_T, Veneto_ ∼ -0.68, *p-value>*0.05), Emilia-Romagna (r_T, Emilia-Romagna_ ∼ -0.55, *p-value<*0.05), Sicilia (r_T, Sicilia_ ∼ -0.39, *p-value<*0.05), Puglia (r_T, Puglia_ ∼ -0.71, *p-value<*0.05). In all Veneto and Emilia-Romagna, the outside weather dropped from 16C to 6C, whereas in Sicilia the temperature dropped from 21C to 13C and in Puglia 20C to 7C. In Veneto and Emilia-Romagna, the cases rose from 1000/cases per day to 2000/cases per day, and in Sicilia and Puglia, the cases rose from 250/cases per day to 1000/cases per day. Both in France and Italy, regions like Grand Est (Fr), Bourgogne-Franche-Comte (Fr), Lombardia (Italy) and Emili-Romagna (Italy), infection rate has weak correlation to relative humidity (r_%RH_ < 0.35, *p-value<0*.*05*).

**Figure 5:**
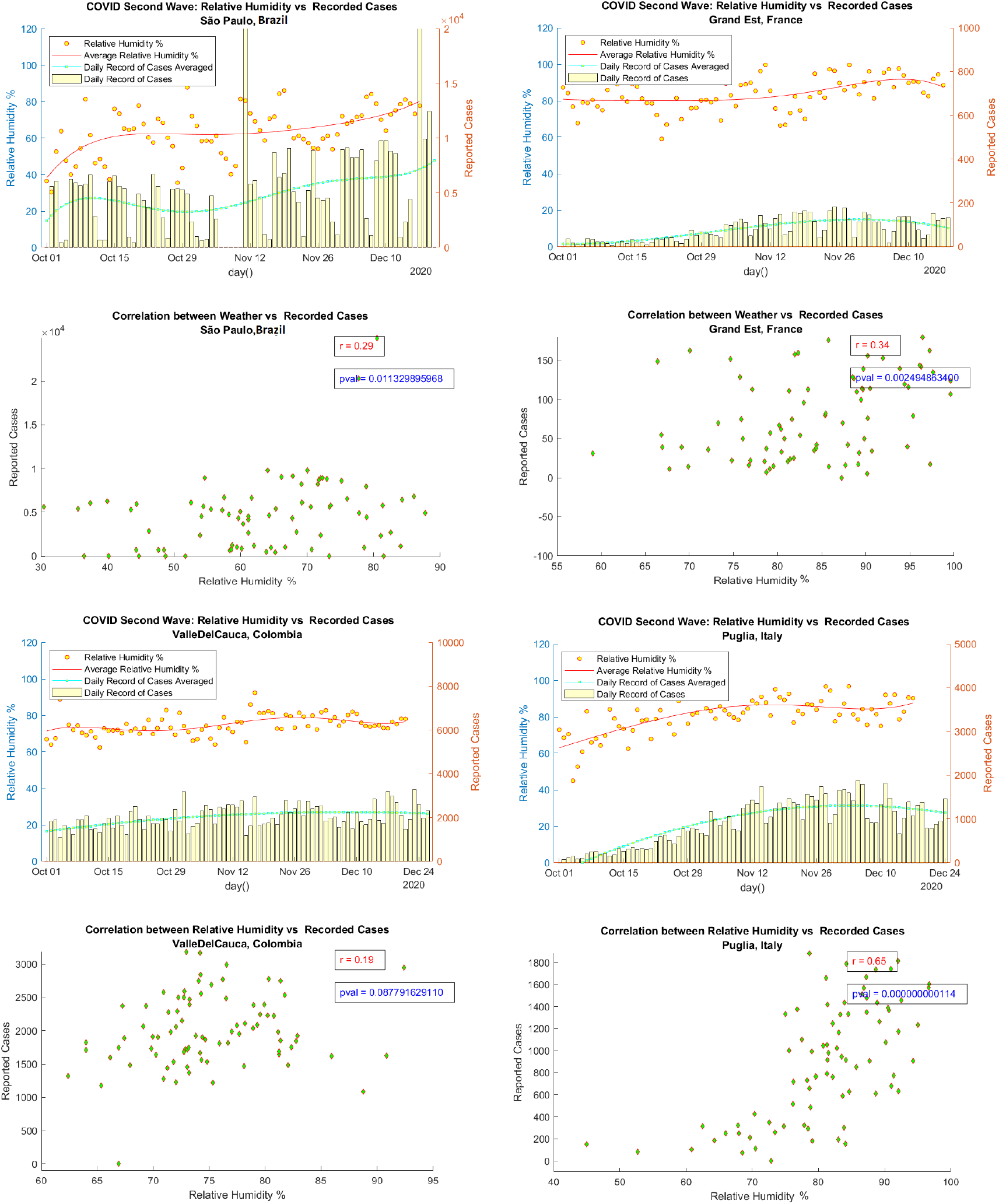
Comparison between relative humidity (%RH) and the COVID infection of four different regions for two different climate types: *São Paulo, Brazil* (r=0.29); *Grand Est, France* (r=0.34); *Valle del Cauca, Colombia* (r=0.19); *Puglia, Italy* (r=0.65)

Even though a thorough study has been made available through this study, a wider look into the dryer climates in middle east, or a varying climatic zone of Australia and colder climates in Canada and Russia could have strengthen the findings and help build a more extensive study on the effect of weather patterns and the spread of COVID infection. A further look into a combination of weather and relative humidity, such like wet-bulb temperature, or other factors like absolute humidity, heat index in dry climate areas should be further explored.

## 4. Conclusion

This study sheds light into the detail of more than 36 regions with widely varying weather patterns. While outside temperature may seem to hold good correlation and might support the hypothesis that outside temperature effects the rate of spread of COVID infection in cold climates such like Italy and France, this hypothesis across warmer humid tropical climates does not hold to be true. With a falling seven-day average outside temperature seemingly causes a rise in infection rate in Italy and France, a very little fluctuation in temperature could not stop the spread of COVID-19 in Colombia. While many of the recent scientific research exploring the strength of correlation between weather and the spread of SARS-CoV-2 may seem to be producing conjectures that are quite convincing, based on this literature findings the notion that COVID-19 is heavily dependable on climate pattern is not convincible and therefore remains quite debatable.

## Data Availability

All data produced in the present study are available upon reasonable request to the authors

## Credit authorship contribution statement

**Ahmed Islam:** Algorithm Development, Resources and Data collection and analysis, Writing - original draft.

## Declaration of competing interest

The authors declare that they have no known competing financial interests or personal relationships that could have appeared to influence the work reported in this paper.

## Notes

### Competing Interest Statement

The authors have declared no competing interest.

### Funding Statement

This study did not receive any funding

