## Supplemental Figures for "The Effect of Weather Pattern on the Second Wave of Coronavirus: A cross study between cold and tropical climates of France, Italy, Colombia, and Brazil"

### Supplemental Information

The information in this page contains report for the four countries and the respective nine most infected regions/territories within the period of October 1$^{st}$ to December 31$^{st}$, 2020.

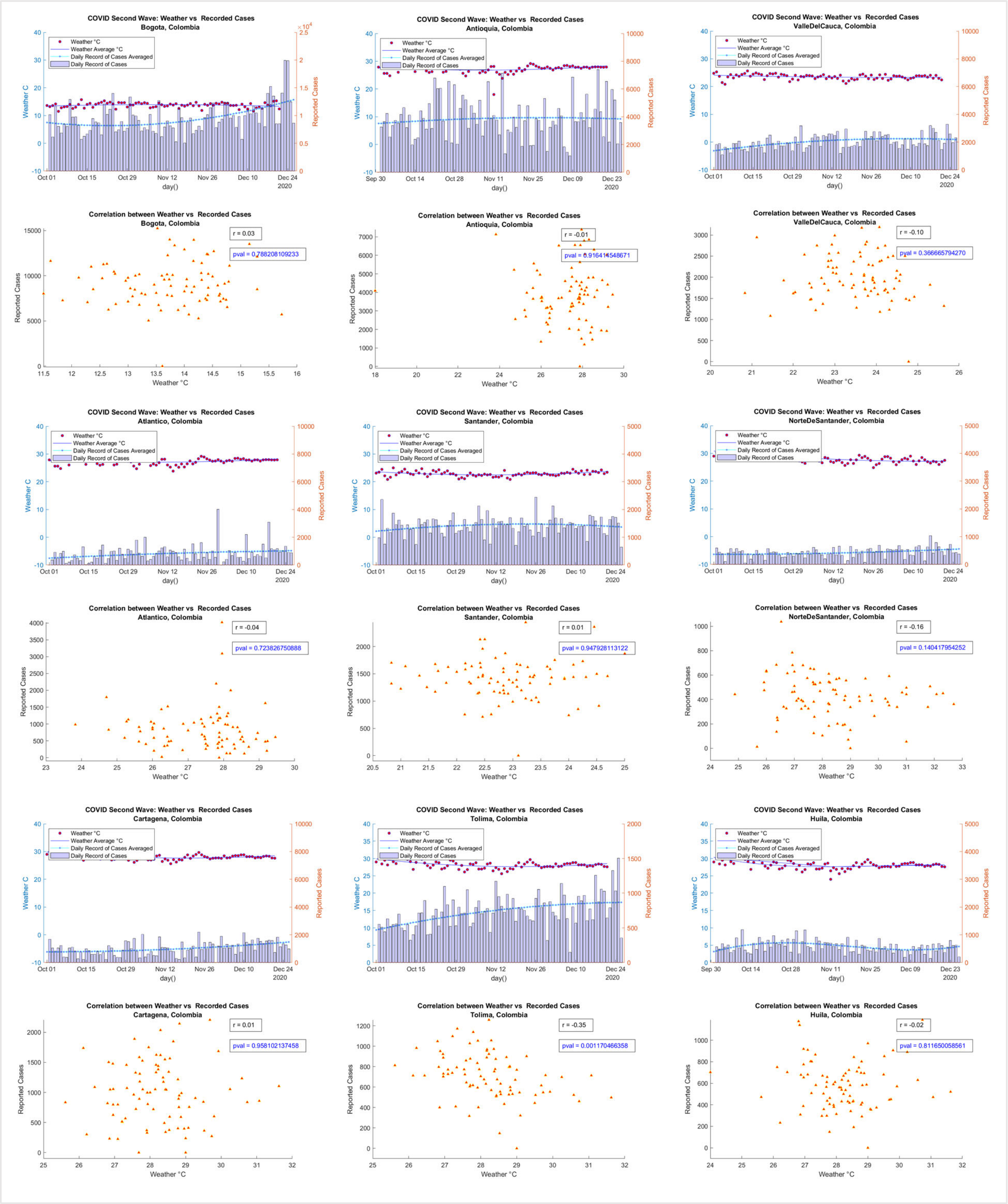

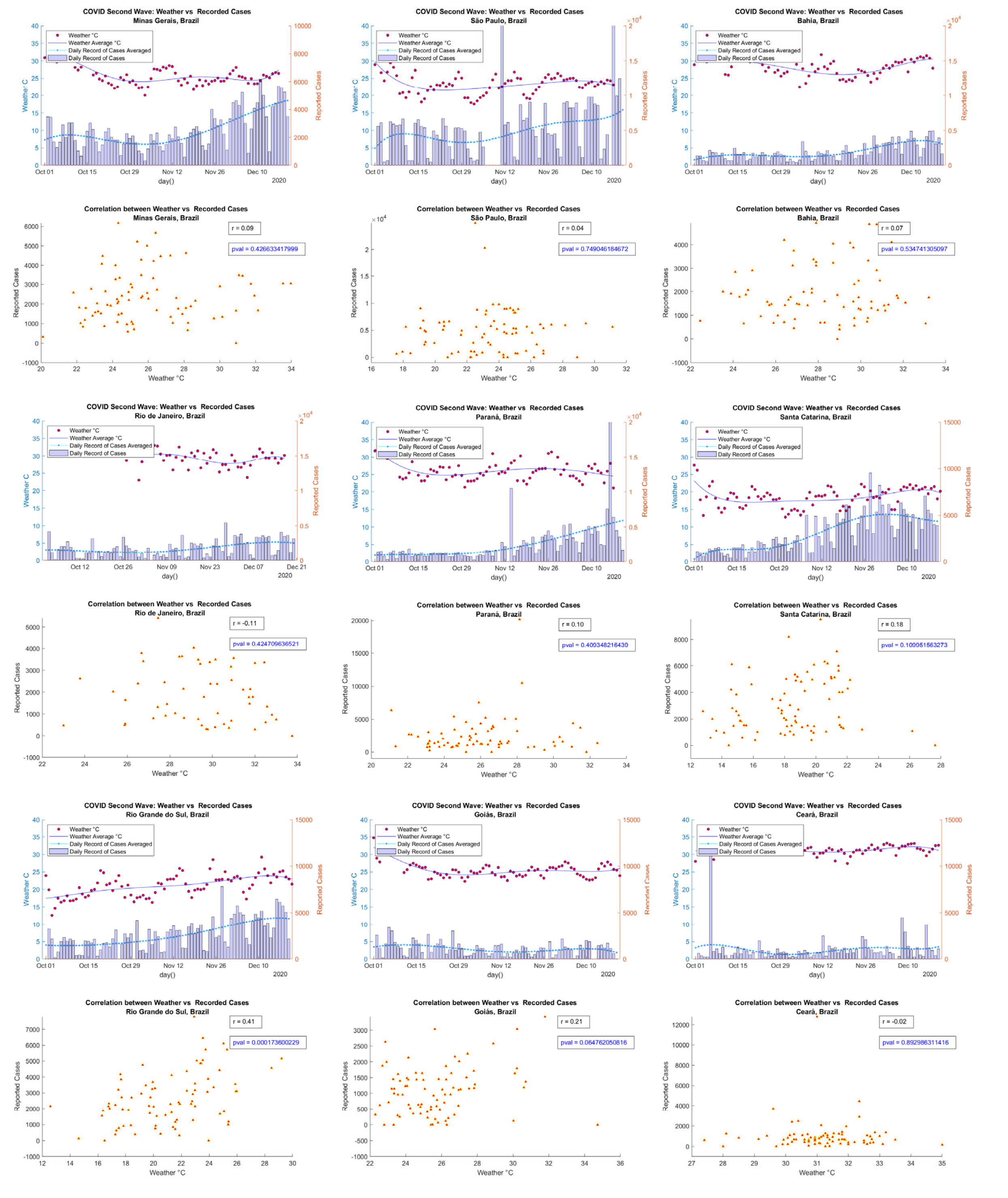
COVID Second Wave: Weather vs Recorded Cases
Minas Gerais, Brazil
COVID Second Wave: Weather vs Recorded Cases
São Paulo, Brazil
COVID Second Wave: Weather vs Recorded Cases
Bahia, Brazil
Weather °C
Weather Average °C
Daily Record of Cases Averaged
Daily Record of Cases
Weather C
Reported Cases
day()
Correlation between Weather vs Recorded Cases
Minas Gerais, Brazil
r = 0.09
pval = 0.426633417999
Correlation between Weather vs Recorded Cases
São Paulo, Brazil
r = 0.04
pval = 0.749046184672
Correlation between Weather vs Recorded Cases
Bahia, Brazil
r = 0.07
pval = 0.534741305097
Weather °C
COVID Second Wave: Weather vs Recorded Cases
Rio de Janeiro, Brazil
COVID Second Wave: Weather vs Recorded Cases
Paranà, Brazil
COVID Second Wave: Weather vs Recorded Cases
Santa Catarina, Brazil
Correlation between Weather vs Recorded Cases
Rio de Janeiro, Brazil
r = -0.11
pval = 0.424709636521
Correlation between Weather vs Recorded Cases
Paranà, Brazil
r = 0.10
pval = 0.400348216430
Correlation between Weather vs Recorded Cases
Santa Catarina, Brazil
r = 0.18
pval = 0.100951563273
COVID Second Wave: Weather vs Recorded Cases
Rio Grande do Sul, Brazil
COVID Second Wave: Weather vs Recorded Cases
Goiás, Brazil
COVID Second Wave: Weather vs Recorded Cases
Cearà, Brazil
Correlation between Weather vs Recorded Cases
Rio Grande do Sul, Brazil
r = 0.41
pval = 0.000173600229
Correlation between Weather vs Recorded Cases
Goiás, Brazil
r = 0.21
pval = 0.064762050816
Correlation between Weather vs Recorded Cases
Cearà, Brazil
r = -0.02
pval = 0.892986311416

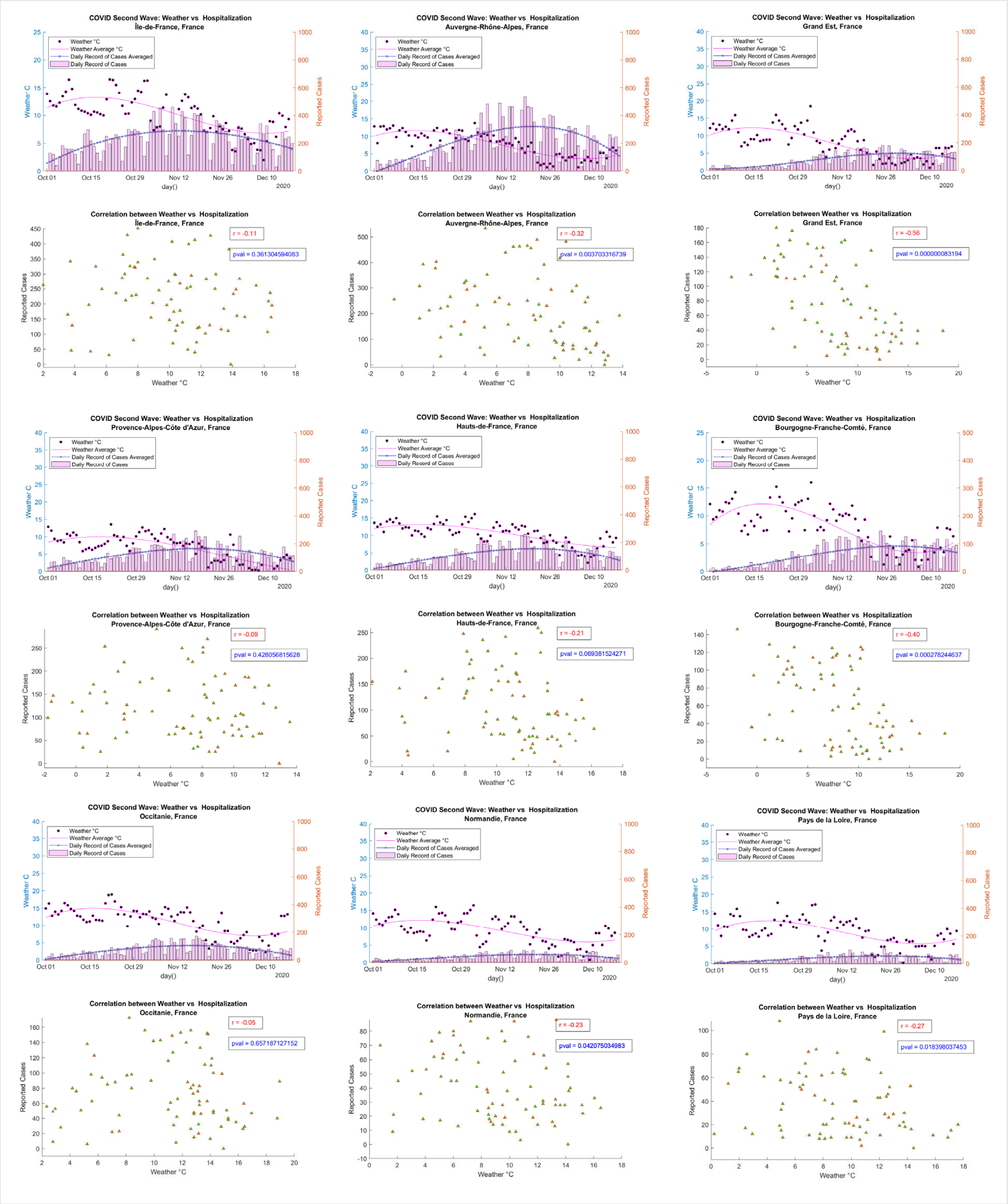
COVID Second Wave: Weather vs Hospitalization
Île-de-France, France
Weather °C
Weather Average °C
Daily Record of Cases Averaged
Daily Record of Cases
Weather C
Reported Cases
day()
Oct 01
Oct 15
Oct 29
Nov 12
Nov 26
Dec 10
2020
COVID Second Wave: Weather vs Hospitalization
Auvergne-Rhône-Alpes, France
COVID Second Wave: Weather vs Hospitalization
Grand Est, France
Correlation between Weather vs Hospitalization
Île-de-France, France
r = -0.11
pval = 0.361304594083
Weather °C
Correlation between Weather vs Hospitalization
Auvergne-Rhône-Alpes, France
r = -0.32
pval = 0.003703316739
Correlation between Weather vs Hospitalization
Grand Est, France
r = -0.56
pval = 0.000000083194
COVID Second Wave: Weather vs Hospitalization
Provence-Alpes-Côte d'Azur, France
COVID Second Wave: Weather vs Hospitalization
Hauts-de-France, France
COVID Second Wave: Weather vs Hospitalization
Bourgogne-Franche-Comté, France
Correlation between Weather vs Hospitalization
Provence-Alpes-Côte d'Azur, France
r = -0.09
pval = 0.428056815628
Correlation between Weather vs Hospitalization
Hauts-de-France, France
r = -0.21
pval = 0.069381524271
Correlation between Weather vs Hospitalization
Bourgogne-Franche-Comté, France
r = -0.40
pval = 0.000278244637
COVID Second Wave: Weather vs Hospitalization
Occitanie, France
COVID Second Wave: Weather vs Hospitalization
Normandie, France
COVID Second Wave: Weather vs Hospitalization
Pays de la Loire, France
Correlation between Weather vs Hospitalization
Occitanie, France
r = -0.05
pval = 0.657187127152
Correlation between Weather vs Hospitalization
Normandie, France
r = -0.23
pval = 0.042075034983
Correlation between Weather vs Hospitalization
Pays de la Loire, France
r = -0.27
pval = 0.018398037453

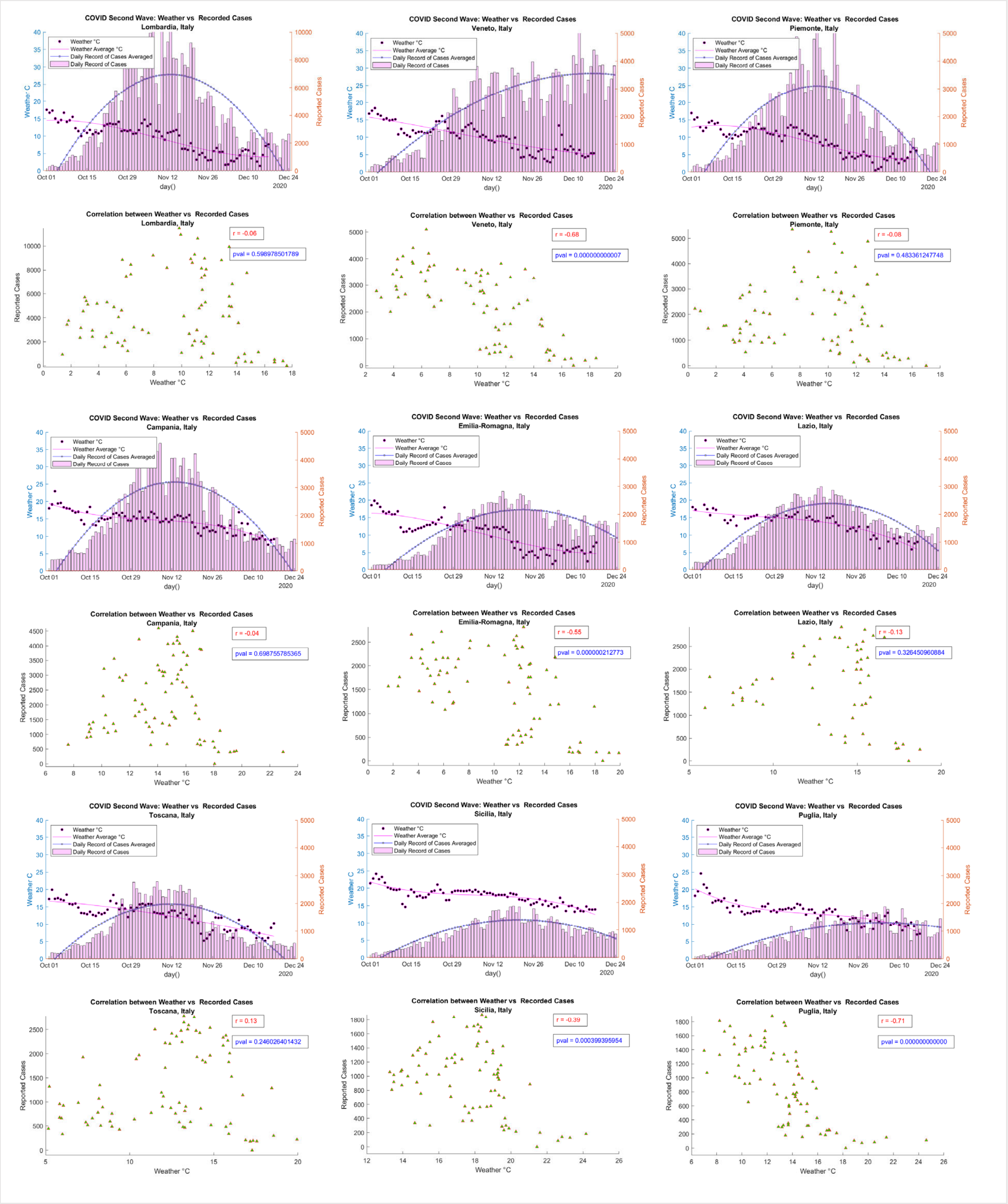
COVID Second Wave: Weather vs Recorded Cases
Lombardia, Italy
Weather °C
Weather Average °C
Daily Record of Cases Averaged
Daily Record of Cases
Weather C
Reported Cases
day()
2020
Correlation between Weather vs Recorded Cases
Lombardia, Italy
r = -0.06
pval = 0.598978501789
Reported Cases
Weather °C
COVID Second Wave: Weather vs Recorded Cases
Veneto, Italy
Correlation between Weather vs Recorded Cases
Veneto, Italy
r = -0.68
pval = 0.000000000007
COVID Second Wave: Weather vs Recorded Cases
Piemonte, Italy
Correlation between Weather vs Recorded Cases
Piemonte, Italy
r = -0.08
pval = 0.483361247748
COVID Second Wave: Weather vs Recorded Cases
Campania, Italy
Correlation between Weather vs Recorded Cases
Campania, Italy
r = -0.04
pval = 0.698755785365
COVID Second Wave: Weather vs Recorded Cases
Emilia-Romagna, Italy
Correlation between Weather vs Recorded Cases
Emilia-Romagna, Italy
r = -0.55
pval = 0.000000212773
COVID Second Wave: Weather vs Recorded Cases
Lazio, Italy
Correlation between Weather vs Recorded Cases
Lazio, Italy
r = -0.13
pval = 0.326450960884
COVID Second Wave: Weather vs Recorded Cases
Toscana, Italy
Correlation between Weather vs Recorded Cases
Toscana, Italy
r = 0.13
pval = 0.246026401432
COVID Second Wave: Weather vs Recorded Cases
Sicilia, Italy
Correlation between Weather vs Recorded Cases
Sicilia, Italy
r = -0.39
pval = 0.000399395954
COVID Second Wave: Weather vs Recorded Cases
Puglia, Italy
Correlation between Weather vs Recorded Cases
Puglia, Italy
r = -0.71
pval = 0.000000000000

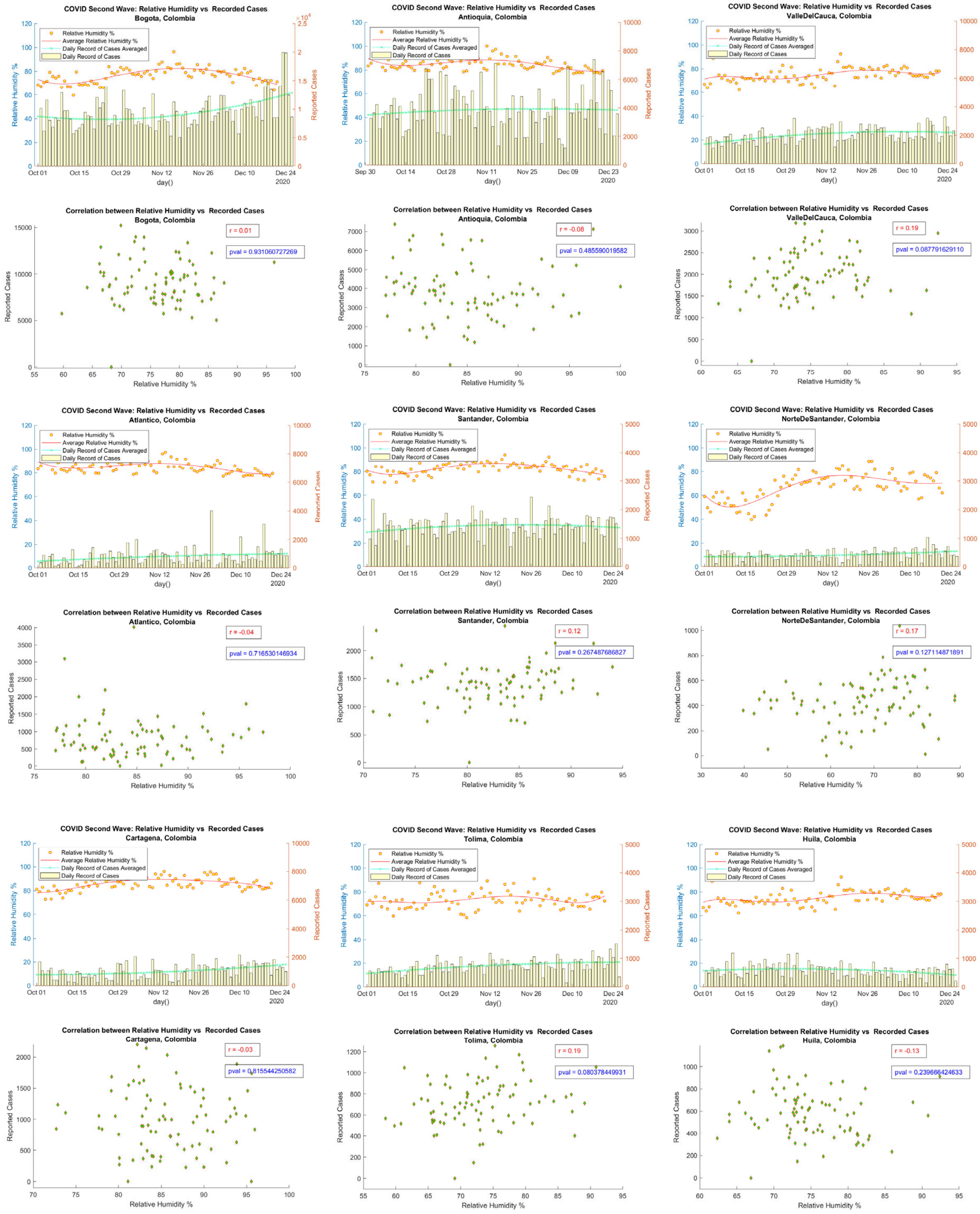

COVID Second Wave: Relative Humidity vs Recorded Cases
Bogota, Colombia
COVID Second Wave: Relative Humidity vs Recorded Cases
Antioquia, Colombia
COVID Second Wave: Relative Humidity vs Recorded Cases
ValleDelCauca, Colombia
Relative Humidity %
Average Relative Humidity %
Daily Record of Cases Averaged
Daily Record of Cases
Relative Humidity %
Reported Cases
day()
Correlation between Relative Humidity vs Recorded Cases
Bogota, Colombia
r = 0.01
pval = 0.931060727269
Correlation between Relative Humidity vs Recorded Cases
Antioquia, Colombia
r = -0.08
pval = 0.485590019582
Correlation between Relative Humidity vs Recorded Cases
ValleDelCauca, Colombia
r = 0.19
pval = 0.087791629110
COVID Second Wave: Relative Humidity vs Recorded Cases
Atlantico, Colombia
COVID Second Wave: Relative Humidity vs Recorded Cases
Santander, Colombia
COVID Second Wave: Relative Humidity vs Recorded Cases
NorteDeSantander, Colombia
Correlation between Relative Humidity vs Recorded Cases
Atlantico, Colombia
r = -0.04
pval = 0.716530146934
Correlation between Relative Humidity vs Recorded Cases
Santander, Colombia
r = 0.12
pval = 0.267487686827
Correlation between Relative Humidity vs Recorded Cases
NorteDeSantander, Colombia
r = 0.17
pval = 0.127114871891
COVID Second Wave: Relative Humidity vs Recorded Cases
Cartagena, Colombia
COVID Second Wave: Relative Humidity vs Recorded Cases
Tolima, Colombia
COVID Second Wave: Relative Humidity vs Recorded Cases
Huila, Colombia
Correlation between Relative Humidity vs Recorded Cases
Cartagena, Colombia
r = -0.03
pval = 0.815544250582
Correlation between Relative Humidity vs Recorded Cases
Tolima, Colombia
r = 0.19
pval = 0.080378449931
Correlation between Relative Humidity vs Recorded Cases
Huila, Colombia
r = -0.13
pval = 0.239666424633

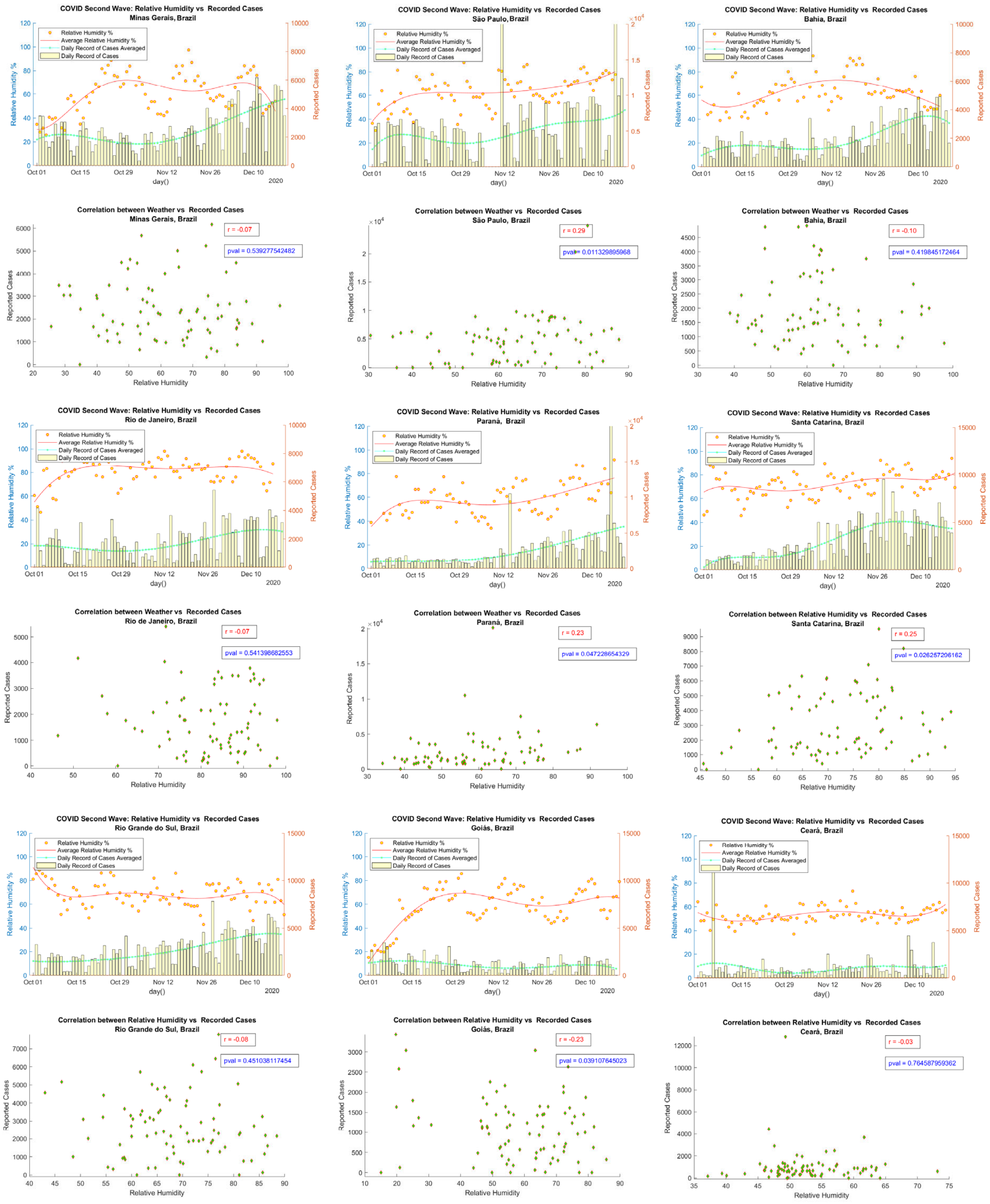
COVID Second Wave: Relative Humidity vs Recorded Cases
Minas Gerais, Brazil
COVID Second Wave: Relative Humidity vs Recorded Cases
São Paulo, Brazil
COVID Second Wave: Relative Humidity vs Recorded Cases
Bahia, Brazil
Relative Humidity %
Average Relative Humidity %
Daily Record of Cases Averaged
Daily Record of Cases
Relative Humidity %
Reported Cases
day()
Oct 01
Oct 15
Oct 29
Nov 12
Nov 26
Dec 10
2020
Correlation between Weather vs Recorded Cases
Minas Gerais, Brazil
r = -0.07
pval = 0.539277542482
Correlation between Weather vs Recorded Cases
São Paulo, Brazil
r = 0.29
pval = 0.011329895968
Correlation between Weather vs Recorded Cases
Bahia, Brazil
r = -0.10
pval = 0.419845172464
Relative Humidity
Reported Cases
COVID Second Wave: Relative Humidity vs Recorded Cases
Rio de Janeiro, Brazil
COVID Second Wave: Relative Humidity vs Recorded Cases
Paraná, Brazil
COVID Second Wave: Relative Humidity vs Recorded Cases
Santa Catarina, Brazil
Correlation between Weather vs Recorded Cases
Rio de Janeiro, Brazil
r = -0.07
pval = 0.541398682553
Correlation between Weather vs Recorded Cases
Paraná, Brazil
r = 0.23
pval = 0.047228654329
Correlation between Relative Humidity vs Recorded Cases
Santa Catarina, Brazil
r = 0.25
pval = 0.026257206162
COVID Second Wave: Relative Humidity vs Recorded Cases
Rio Grande do Sul, Brazil
COVID Second Wave: Relative Humidity vs Recorded Cases
Goiás, Brazil
COVID Second Wave: Relative Humidity vs Recorded Cases
Ceará, Brazil
Correlation between Relative Humidity vs Recorded Cases
Rio Grande do Sul, Brazil
r = -0.08
pval = 0.451038117454
Correlation between Relative Humidity vs Recorded Cases
Goiás, Brazil
r = -0.23
pval = 0.039107645023
Correlation between Relative Humidity vs Recorded Cases
Ceará, Brazil
r = -0.03
pval = 0.764587959362

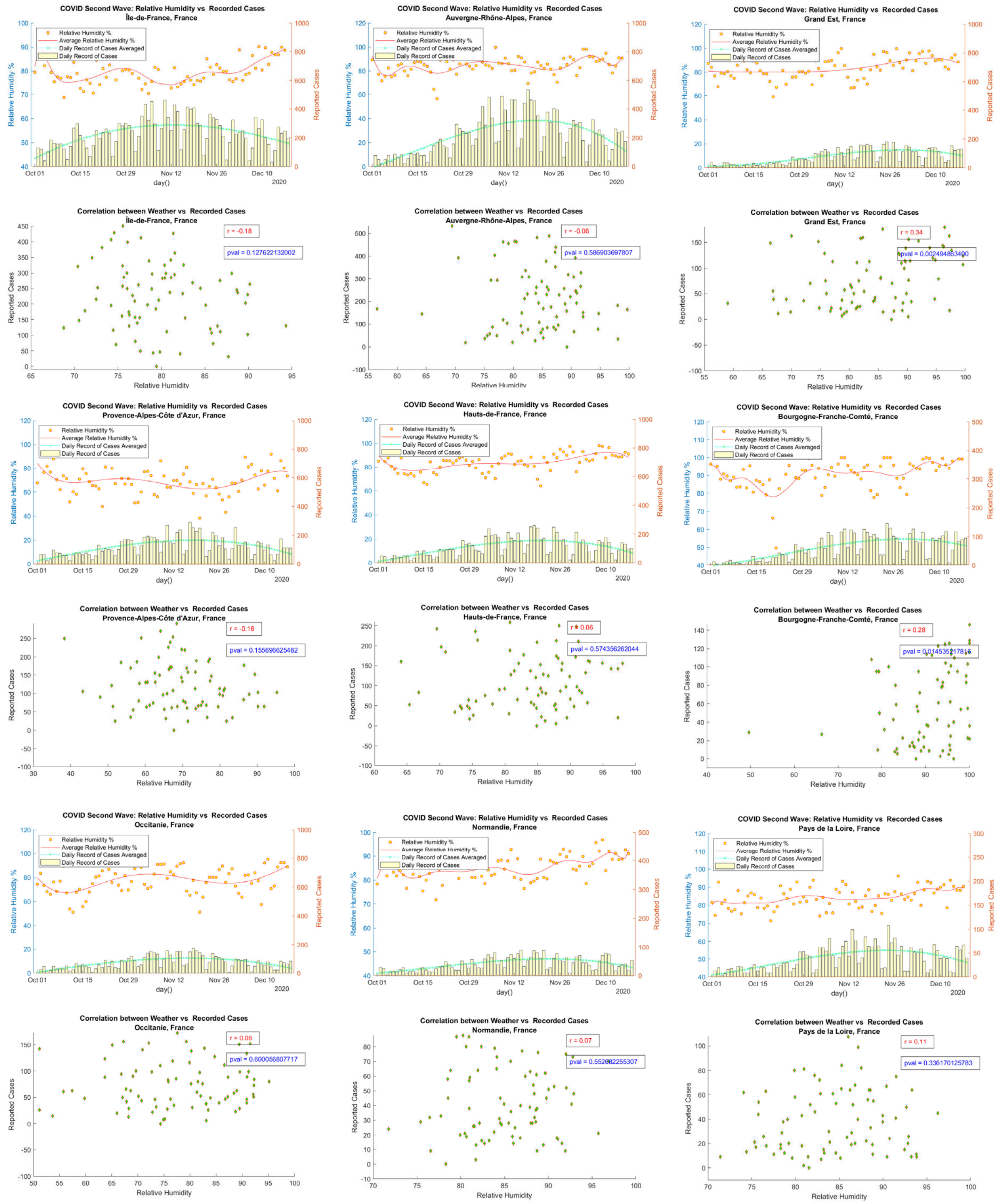
COVID Second Wave: Relative Humidity vs Recorded Cases
Île-de-France, France
Correlation between Weather vs Recorded Cases
Île-de-France, France
r = -0.18
pval = 0.127622132002
COVID Second Wave: Relative Humidity vs Recorded Cases
Auvergne-Rhône-Alpes, France
Correlation between Weather vs Recorded Cases
Auvergne-Rhône-Alpes, France
r = -0.06
pval = 0.586903697807
COVID Second Wave: Relative Humidity vs Recorded Cases
Grand Est, France
Correlation between Weather vs Recorded Cases
Grand Est, France
r = 0.34
pval = 0.002494863400
COVID Second Wave: Relative Humidity vs Recorded Cases
Provence-Alpes-Côte d'Azur, France
Correlation between Weather vs Recorded Cases
Provence-Alpes-Côte d'Azur, France
r = -0.16
pval = 0.155696625482
COVID Second Wave: Relative Humidity vs Recorded Cases
Hauts-de-France, France
Correlation between Weather vs Recorded Cases
Hauts-de-France, France
r = 0.06
pval = 0.574356262044
COVID Second Wave: Relative Humidity vs Recorded Cases
Bourgogne-Franche-Comté, France
Correlation between Weather vs Recorded Cases
Bourgogne-Franche-Comté, France
r = 0.28
pval = 0.014535217816
COVID Second Wave: Relative Humidity vs Recorded Cases
Occitanie, France
Correlation between Weather vs Recorded Cases
Occitanie, France
r = 0.06
pval = 0.600056807717
COVID Second Wave: Relative Humidity vs Recorded Cases
Normandie, France
Correlation between Weather vs Recorded Cases
Normandie, France
r = 0.07
COVID Second Wave: Relative Humidity vs Recorded Cases
Pays de la Loire, France
Correlation between Weather vs Recorded Cases
Pays de la Loire, France
r = 0.11
pval = 0.336170125783
Relative Humidity %
Average Relative Humidity %
Daily Record of Cases Averaged
Daily Record of Cases
Relative Humidity
Reported Cases
day()

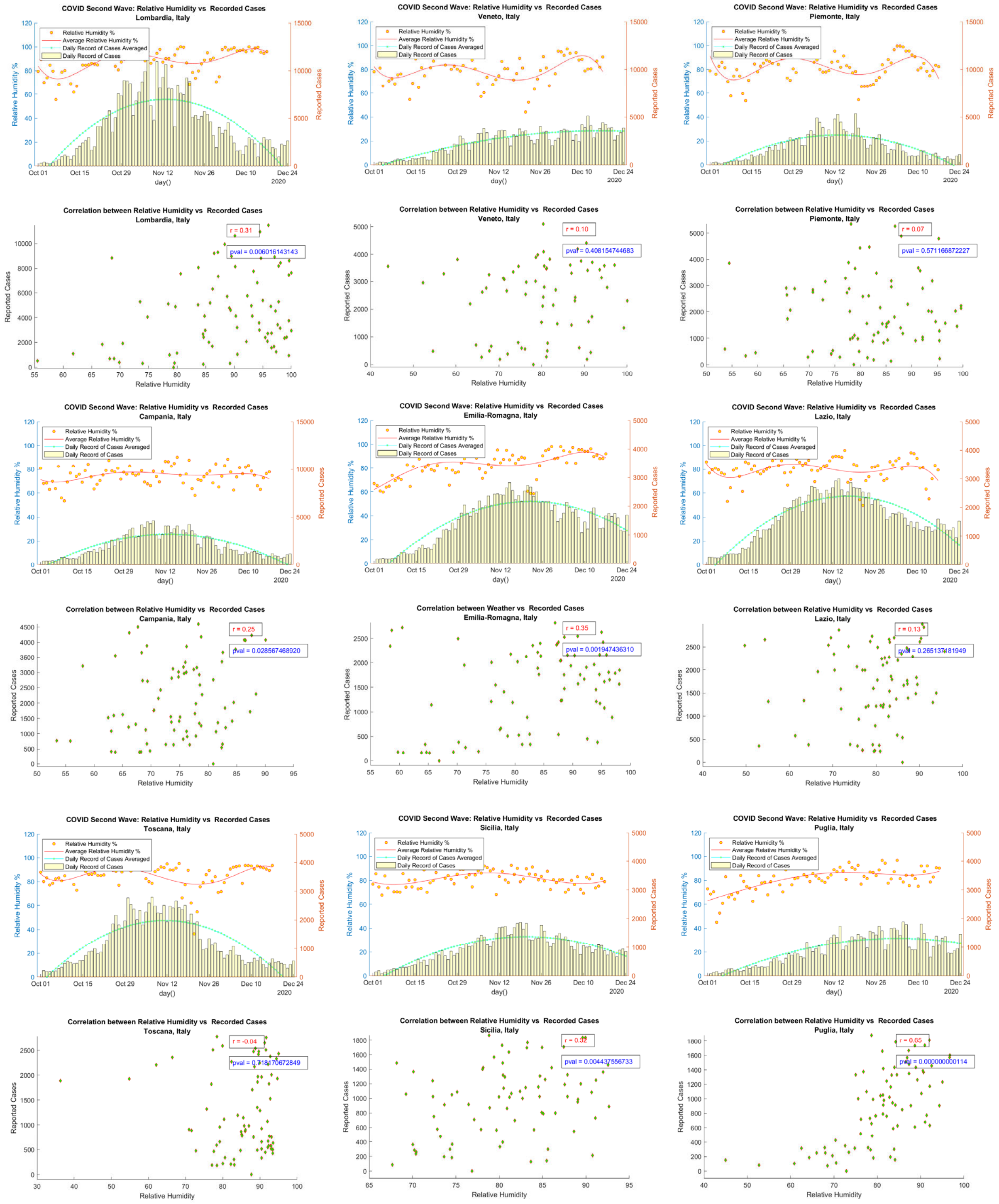

COVID Second Wave: Relative Humidity vs Recorded Cases
Lombardia, Italy
Relative Humidity %
Average Relative Humidity %
Daily Record of Cases Averaged
Daily Record of Cases
Relative Humidity %
Reported Cases
day()
2020
COVID Second Wave: Relative Humidity vs Recorded Cases
Veneto, Italy
COVID Second Wave: Relative Humidity vs Recorded Cases
Piemonte, Italy
Correlation between Relative Humidity vs Recorded Cases
Lombardia, Italy
r = 0.31
pval = 0.006016143143
Correlation between Relative Humidity vs Recorded Cases
Veneto, Italy
r = 0.10
pval = 0.408154744683
Correlation between Relative Humidity vs Recorded Cases
Piemonte, Italy
r = 0.07
pval = 0.571166872227
Relative Humidity
COVID Second Wave: Relative Humidity vs Recorded Cases
Campania, Italy
COVID Second Wave: Relative Humidity vs Recorded Cases
Emilia-Romagna, Italy
COVID Second Wave: Relative Humidity vs Recorded Cases
Lazio, Italy
Correlation between Relative Humidity vs Recorded Cases
Campania, Italy
r = 0.25
pval = 0.028567468920
Correlation between Weather vs Recorded Cases
Emilia-Romagna, Italy
r = 0.35
pval = 0.001947436310
Correlation between Relative Humidity vs Recorded Cases
Lazio, Italy
r = 0.13
pval = 0.265137481949
COVID Second Wave: Relative Humidity vs Recorded Cases
Toscana, Italy
COVID Second Wave: Relative Humidity vs Recorded Cases
Sicilia, Italy
COVID Second Wave: Relative Humidity vs Recorded Cases
Puglia, Italy
Correlation between Relative Humidity vs Recorded Cases
Toscana, Italy
r = -0.04
pval = 0.718170672849
Correlation between Relative Humidity vs Recorded Cases
Sicilia, Italy
r = 0.32
pval = 0.004437556733
Correlation between Relative Humidity vs Recorded Cases
Puglia, Italy
r = 0.65
pval = 0.000000000114
